# ‘Multisectoral socialisation’ in national health governance: An ethnographic study of Singapore’s One Health approach

**DOI:** 10.64898/2026.01.12.26343975

**Authors:** Matthew Aviso, Youming Ng, James Chan, Panqin Cai, Kelvin Lim, Natasha Howard

## Abstract

**Introduction:** The 2025 establishment of Singapore’s Communicable Diseases Agency (CDA) as the country’s national public health agency included creation of a One Health Office (OHO). OHO’s establishment supports the pre-existing One Health whole-of-government approach, consisting of multisectoral collaborations via five government agencies spanning human, animal, and environment-related health. Singapore’s One Health approach includes several unique features designed to stimulate preparedness, prevention, and mitigation measures during periods of non-emergency. We thus aimed to analyse how this approach developed and implemented multisectoral governance of national One Health priorities.

**Methods:** This ethnographic study, conducted over 9 months (2024-2025), included unstructured observations of 3 inter-agency One Health meetings and 8 partner agency engagements, 26 key informant interviews with public servants across the five One Health partner agencies, and review of 8 internal government documents (e.g. draft plans, reports, presentation slides). We analysed data thematically using abductive coding.

**Findings:** Foundational to Singapore’s One Health governance approach, we found a participant-driven socialisation process dependent on the development of ‘semi-informal’ inter-agency relations among public servants. A rotational governance structure and small size of the city-state fostered these relations, which in turn strengthened/were strengthened by regular bilateral and multilateral communication and engagement by OHO to achieve workable arrangements while continuously building multisectoral governance of One Health nationally.

**Conclusions:** Singapore’s One Health approach and emergent multisectoral governance is predicated on cultivating interpersonal socialisation among stakeholders. Agencies such as CDA exemplify how formal structures can be strengthened through “semi-informal” social networks that foster shared ownership, trust, and consensus across government entities. For national public health agencies elsewhere, embedding such socialisation processes within governance may enhance collaboration and ownership building, even across multiple sectors.

## INTRODUCTION

Multisectoral governance refers to coordination and collaboration among several sectors (e.g. health, agriculture, environment, urban planning) to address complex challenges that span disciplinary and organisational boundaries, emphasising collective decision-making, shared responsibility, and integrated responses to achieve common goals [1,2]. The city-state of Singapore has intentionally developed multisectoral partnerships for health governance, with Singapore’s successful COVID-19 response demonstrating the value of multisectoral and ‘whole-of-government’ (WOG) approaches. The 2023 COVID-19 White Paper proposed strengthening these approaches for pandemic planning and preparedness, given the increasing frequency of infectious disease outbreaks and antimicrobial resistance [3]. Spurred by noted gaps in inter-agency partnerships in the national COVID-19 pandemic response, Singapore established an interim Communicable Diseases Agency (iCDA) in 2024—a pre-operational model to facilitate transition into the Communicable Diseases Agency (CDA) [4]. CDA became operational in April 2025 as a national public health agency (NPHA) that functions as statutory board under the Ministry of Health (MOH) and consolidates public health functions previously spread across the Ministry of Health (MOH), National Centre for Infectious Diseases (NCID), and Health Promotion Board (HPB) [4].

CDA thus functions both as part of and autonomously from MOH, serving as the frontline government agency to prevent, prepare for, detect, quickly respond to, and enable research in infectious diseases in Singapore. This includes leading and coordinating public health preparedness; safeguarding national interests such as vaccine and therapeutics development; strengthening surveillance capabilities through new modalities such as wastewater testing, data analytics, and artificial intelligence; and conducting contact tracing and border control measures [5]. Minister of State for Health Rahayu Mahzam indicated that MOH will drive Singapore’s pandemic approach and strategy, while CDA will provide policy and scientific recommendations [4] as part of ongoing multisectoral approaches to addressing current and emerging public health threats. While CDA is still in its infancy, it is helpful to examine pre-existing multisectoral health governance efforts in the country to inform governance models for CDA and other NPHAs. This study thus focuses on the new One Health Office (OHO), a CDA division, to explore the development of multisectoral governance approaches in Singapore.

The concept of “One Health” as a collaborative and integrative approach to the health of humans, animals, and ecosystems was adopted by the World Health Organization (WHO) in 2008 [6]. Multisectoral coordination is a foundational principle of the One Health approach, emphasising inclusivity, accountability, and coordination among diverse human and animal health stakeholders, to improve decision-making and resource usage in addressing human, animal, and ecologically related disease threats [7]. From avian influenza to Ebola and, more recently, mpox, linkages between humans, other animals, and the environment require multisectoral action to prevent and mitigate emergencies [8–11].

Recognizing this need for multisectoral collaboration, Singapore established a formal inter-agency One Health framework in 2012, including MOH (now through CDA), the National Environment Agency (NEA) to mitigate impacts of vector-borne diseases such as dengue fever, Singapore Food Agency (SFA) for food safety, National Parks Board (NParks) for diseases that may affect both animals and humans, and Singapore’s National Water Agency (PUB) for water safety. This initiative seeks to address emergent global health challenges in an era of climate and ecosystem changes that have displaced multiple animal species, fostering the rapid development of microbial beings that, in the worst case, become pandemic-level threats [12]. Significant progress included developing joint response protocols for priority diseases, strengthening workforce capacity, and establishing risk communication and surveillance workflows [13, 14].

The COVID-19 pandemic prompted Singapore to strengthen One Health measures and re-envision this approach, leading to OHO’s establishment in early 2024 as part of the then-iCDA. Under CDA, OHO aims to bolster intersectoral governance, policy, and advocacy development. Since its inception, OHO has coordinated stakeholders from the five national lead agencies to develop strategic direction and measurable actions for the One Health space. Towards that end, it had been developing a One Health master plan—a national strategy to safeguard public health, representing the culmination of collaborative efforts across Singapore’s health, environmental, and animal sectors, and intended to be a living document to incorporate stakeholder feedback over time. These documents are developed based on multiple consultation and consensus-building inter-agency meetings attended by human disease control professionals, veterinarians, scientists, wildlife ecologists, or those working in related disciplines. Via these activities, CDA intends to establish itself through OHO as a coordinating body for multisectoral One Health collaborations across partner agencies and external stakeholders.

We aimed to analyse how the OHO and Singapore’s One Health approach develop and implement multisectoral governance of national One Health priorities. Objectives were to: (i) describe the structural processes through which the One Health approach fosters multisectoral governance efforts among different agencies; (ii) explore how inter-agency One Health meetings demonstrate multisectoral governance processes; and (iii) identify key lessons that can inform national and global multisectoral governance of NPHAs.

## METHODOLOGY

### Study design

We adopted an ethnographic study design [15, 16], drawing from descriptive and exploratory data gathered from semi-structured interviews [17], participant observations, and document review. Ethnography is a fieldwork methodology that attempts to collect “thick” descriptive data in the form of observational fieldnotes to describe the “insider” perspective of a social or cultural group. It is particularly suited for collecting detailed qualitative data related to social interactions and processes and involves reflexively negotiating the impact of one’s “outsider” presence on the proceedings of the observations [18]. Rather than coming from a position of objectivity, ethnography acknowledges that all qualitative data are inherently subjective and cannot be elicited or extracted in a controlled setting as laboratory data can. Therefore, in contrast to a methodologically similar study conducted on health policy spaces in Singapore that considers itself as engaging in “non-participant observation” [19], we consider ourselves participant observers whose presence cannot be divorced from the field within which we make our observations. Our research question was: “How does Singapore’s One Health approach help develop or strengthen multisectoral governance?”

### Data collection

#### Participant observation

MA observed a series of OHO-coordinated inter-agency workshops and meetings to describe the interactions and activities occurring in these spaces. MA prepared ethnographic fieldnotes of proceedings, including observations and conversations among attendees. Semi-structured observations (e.g. of verbal intonation and body language; affirmative or disconfirming assertions; acts of disengagement, challenge, or submission) and ethnographic conversations were noted through traditional fieldnote-taking. Ethnographic conversations are unstructured and iterative by nature and thus did not follow a pre-prepared question guide, instead documenting and clarifying important or interesting points that may not be obtained through observations or formal semi-structured interviews. These conversations arose spontaneously during meetings, were not audio-recorded, and were used to inform interpretations of observations and revise lines of questioning in the semi-structured interview guide.

From 1 September 2024 to 31 June 2025, MA attended two One Health scoping workshops for masterplan development and one meeting for action plan drafting, with eight site visits to representative agency offices where observational notes were taken alongside in-person interview notes. OHO and partner agencies provided permission to attend meetings or visit offices for observation and conversation, with MA’s meeting attendance limited to multisectoral scoping meetings. Each OHO-coordinated event MA attended lasted three to seven hours.

#### Key informant interviews

We developed a question guide based on multisectoral governance literature, observations, and document review. It included topics on the national One Health governance approach, institutional and personal relationship with CDA and OHO, limitations and possibilities of inter-agency One Health work, shared experiences of inter-agency gatherings—where MA clarified participant thoughts on key moments of the meetings to also corroborate observation fieldnotes—and experiences of meetings MA did not attend. These last constituted a form of oral history often employed in ethnographic approaches [20].

We used purposive maximum variation sampling [21, 22] as the most feasible way to sample across the small pool of 65 candidate public servants, based on a list of attendees in inter-agency One Health gatherings since late 2024. Representation from across the five agencies in these meetings are roughly equal in terms of attendee count. Eligibility included public servants who: (i) had worked in Singapore public service for a cumulative three years prior to interview, and (ii) attended at least one inter-agency One Health gathering as participant or facilitator between 2024 and 2025. Public servants were initially approached in-person at One Health meetings. Due to the dynamic nature of public service, scheduling constraints limited the number of initial responses. We then employed snowballing, whereby participants were asked to recommend other participants who we contacted via email or in-person during succeeding One Health gatherings. This facilitated the identification of candidates and inclusion of individuals with specialised knowledge or unique perspectives that may otherwise have been overlooked. We strove to proportionately represent interviewees across different ranks while also balancing representation from each agency. Doing so was an attempt to maximise variation of ideas and backgrounds in a small population size.

We provided potential interviewees with the study information sheet and opportunities to ask questions before recording written or verbal (as participant’s preferred) consent before interviews. MA conducted all interviews between March-May 2025, at places of participants’ choosing (typically their offices) or remotely using Zoom (Zoom Video Communications Inc, San Jose, California) or Microsoft Teams software. Interviews were audio-recorded through Zoom or a separate audio-recording device, with automatic transcription enabled. One participant was interviewed through writing per their request, while five did not wish to be recorded but allowed notetaking. Interviews lasted 45 minutes on average, with MA reviewing and correcting transcripts after each interview.

#### Document review

We reviewed 8 internal documents provided by OHO, to provide contextual understanding of interviews and observations. These included slide decks presented in meetings the team was not invited to attend, summaries of proceedings, participant lists, updated terms of reference, and drafts of One Health planning documents.

### Analysis

MA analysed data in NVivo 11 software (Lumivero LLC) employing Braun and Clarke’s thematic analysis [23] with an abductive approach described by Thompson [24, 25]. This integrated deductive coding based on the question guide while leaving analysis open to new insights not covered by initial questions. This included in-depth (re)reading of transcripts, initial open coding, refinement into broad themes, recombination, and interpretation/contextualisation. Interpretation and contextualisation were aided by review of internal OHO documents. An abductive approach was most appropriate as we began with a set of questions to explore and explain specific structural features of the One Health space but yielded new ideas related to socialisation processes. Abductive analysis is common in ethnography, as initial assumptions and questions change and develop through the course of fieldwork. Analysis was conducted simultaneously with data collection. Later interviews thus included questions and discussions touching on tentative thematic analysis, constituting a form of participant checking.

### Ethics

The National University of Singapore Institutional Review Board provided ethics approval (reference NUS-IRB-2025-41).

## FINDINGS

### Participant characteristics and themes

We interviewed 26 participants, 18 individually and 8 in three group interviews, with proportional representation across the different One Health structure levels after CDA formation (Figure 1). In this structure, staff-level participants at director-level and below were asked by OHO for inputs on the One Health masterplan and action plan, and these same members were organized to develop action items. The secretariat served as focal point through which programmes and communications were coordinated from OHO to relevant stakeholders in their home agencies. OHO functioned by design as secretariat for the CDA. The One Health Working Group consisted of members from senior management (i.e. Group Director-equivalent) who developed and coordinated programmes and activities based on staff-level input. The One Health Coordinating Committee (OHCC) oversaw the One Health space, consisting of members of senior management (i.e. Director General-equivalent) that review and endorse programmes and activities proposed by the OHWG and make final decisions based on consensus.

**Figure 1.**
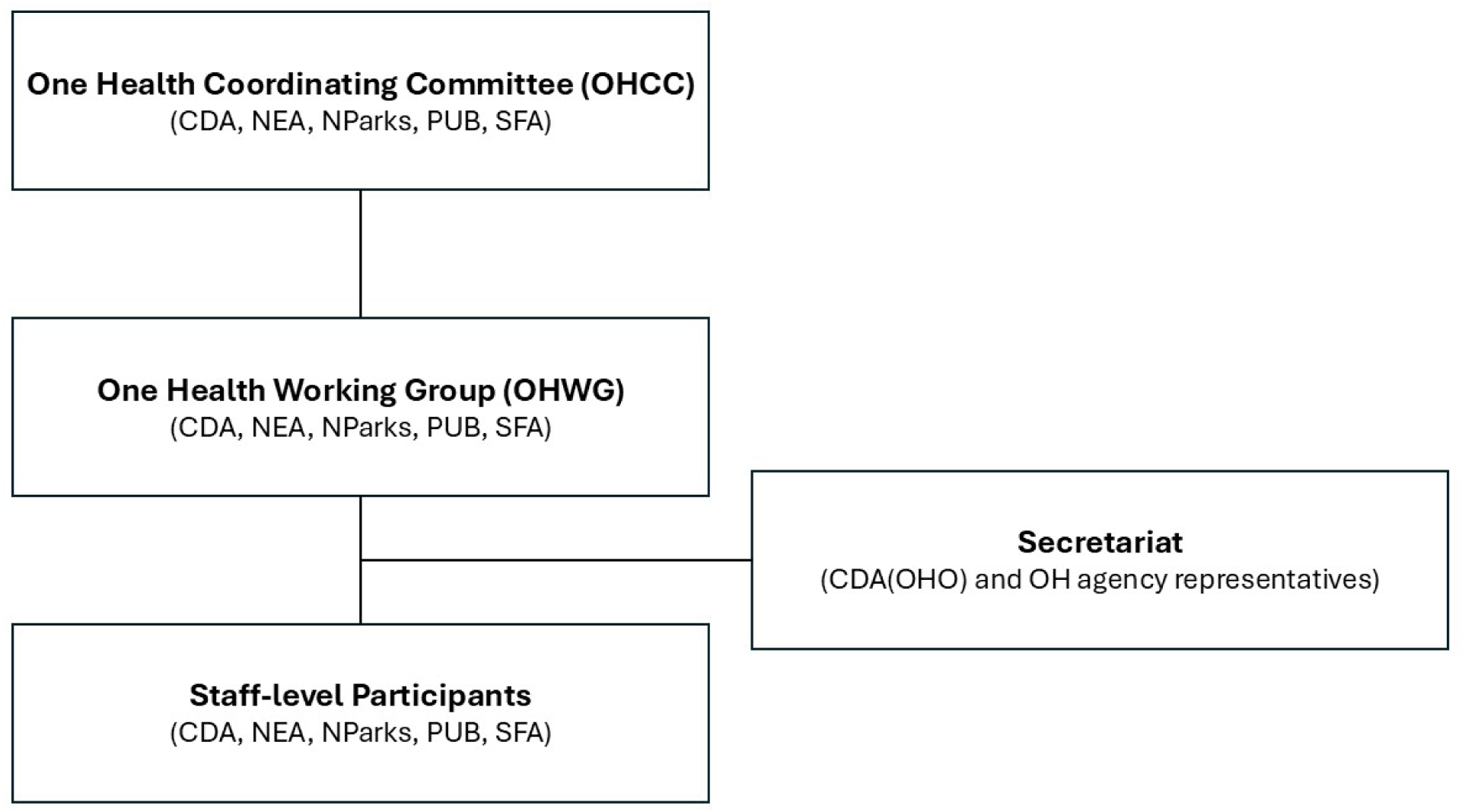
Singapore One Health structure (as of August 2025)

Figure 1 presents Singapore’s One Health structure during the period of data collection, with all participant insights and opinions based on experiences with this structure.

Relatively equal numbers of interviewees across agencies were engaged, with specific numbers not provided to protect the confidentiality of the small number of people from each agency. Several junior or staff-level participants were previously engaged in committees drafting the One Health masterplan and action plan. We also limited details on agency and other identifying details to help ensure anonymity in this small community. Five potential interviewees declined participation, instead connecting us with other One Health stakeholders to interview.

We generated four abductive themes: (i) distributing equal ownership through rotating chairmanship; (ii) centring communicable human diseases; (iii) fostering ‘semi-informal’ relations; and (iv) deepening connections through coordinated bilateral engagement. Findings below reflect participants’ perceptions of Singapore’s One Health approach.

### Distributing equal ownership through rotating chairmanship

Rotating chairmanship was a structural feature of the Singapore One Health approach. This meant that different agencies took turns annually to chair meetings and set agendas. Even before OHO formation, rotating chairmanship was considered a “signal of neutrality.” This was deemed particularly important for non-crisis work, as opposed to just letting one agency such as MOH (or now, CDA) take the lead, as argued by an interviewee:

> Having a lead is great for crisis when there are a lot of disagreement because somebody will decide and everybody falls in line. But at the same time if you only have one agency, the other agency might say, ∜Hey, I’m not responsible.” So, it’s the other agency’s responsibility. Would they scale down some of their involvement? But because it’s rotating, everybody has a responsibility.

This culture of rotation across levels and agencies, however, is not new in Singapore public service. For junior-level officers, rotations allow them to be exposed to different areas of work within their home agency or others. For instance, within agencies, the secretariat rotates among junior staff, though specifics may differ by agency. As a participant described:

> People movement is important, and I’m not sure whether other countries do that [for junior staff], where they have cross posting of people […] For us, it’s for grooming and development but it’s also for understanding the other agencies as well.

Even senior management are rotated frequently in Singapore public service. Such a rotational system was affirmed by many participants as a strength in national governance structures, as transferring diverse technical experts around different portfolios in different agencies helped the cross-fertilisation of ideas—a feature that is especially important for One Health, which is collaborative and complex by design. One senior official advocated for this model particularly strongly:

> Something that I’ve noticed that is quite unique in Singapore…and something I have also recommended and pushed quite a lot internationally is how common[ly] staff and people get rotated. Yeah, I think that it’s very common [that] medical doctors are inside in MOH. Then the veterinarians are in other agencies […but now there’s] a vet [seconded] to MOH to do One Health. And there are many of this going on, especially when it comes to leadership. Our CEO will move. So it can be very tiring because you keep changing CEO. But as I see the system, as I grow, as I grow older, I see the power of this movement […These CEOs] also bring our agenda to the next ministry [they handle]. So there is a lot of mutual understanding between agencies because of this movement of people. If you stay put in one area for too long—twenty, thirty years—you become too guarded.

Still, some acknowledged that rotating chairmanship could have unintended consequences for perceived responsibility in One Health, with one participant noting:

> It’s always circled back to the discussion that it’s an equal ownership of One Health and therefore we do the rotating thing. But if everyone owns it equally, sometimes nobody owns it because, everyone’s kind of like…. it’s all our jobs. You know what I mean? Like if you don’t put someone and say that they’re the ones in charge, then it’s hard to get together a concerted effort.

OHO formation sought to resolve the issue of dispersed rotational efforts by centralising the bulk of coordination, administration, and documentation work while still ensuring each agency took turns annually to chair activities and table One Health items. The OHO functions as central secretariat, with a senior official describing its presence as making sure “there’s some longevity and drive to maybe some of the major initiatives that the agencies may want to pursue,” although it will not necessarily drive technical leadership and expertise.

### Centring communicable human diseases

OHO was young and its secretariat staff strove to demonstrate in meetings that they were coordinators and not just there to provide the CDA (i.e. human infectious diseases-centred) perspective. One participant observed:

> At least from an optics point of view, One Health Office makes a point to distinguish themselves from [the rest of the CDA] in the sense that when they run a meeting that’s just the Secretariat, they don’t offer the opinions as a One Health body.

Nonetheless, many participants indicated that the CDA should house OHO as it is only appropriate that a human health-centred entity should be the structural centre of Singapore’s One Health space. As one interviewee noted:

> I personally think it’s correct to be under MOH [via its statutory board CDA], all right… As an urban city, it is really human density that is high. So, if there’s any disease transmission, it will be human. So, I think from the, especially when it comes to the Singapore perspective, [One Health being centralised] under MOH is correct.

However, some also expressed concerns that focusing on infectious diseases in humans and pandemic preparedness could limit discussion of other important One Health issues.

> Just one possibility, because it’s under the Communicable Diseases Agency […] there may be other One Health threats that are not communicable. So […], if it becomes a need, [we need to discuss] whether that would be something that CDA is comfortable with.

For instance, one participant wanted to centre chemical pollution’s impact on human health in future One Health meetings:

> Environmental pollutants or contaminants that are starting to affect human health as well because of our exposure to them […] These are also one health areas. But I don’t think that we have been talking about these enough. I think we’ve only been talking about viruses and bacteria causing diseases […] I feel that, you know, for Singapore, maybe we also want to have a conversation about… do we also start to look at these other areas besides the conventional bacteria and viruses?

Another interviewee was optimistic that the institutional structures in place and the relationships these fostered would make discussion regarding such grey areas within national One Health priorities fruitful:

> I think with CDA coming in, we have very much scope to look into more of the infectious disease space. But given that we have about 10 years of working together, even though it is not infectious disease, [even if] it could be a chemical hazard that might involve a couple of the agencies, I think that that familiarity and that way of talking and working does come in even for things that might now fall outside the [human infection-centred] One Health.

### Fostering semi-informal relations

Many participants described the abovementioned “familiarity” and relative ease with which they could connect with others as characteristic of a “semi-informal” relationship—described as being able to maintain professional boundaries while still being friendly and accessible to each other. Most participants suggested that such semi-informal relationships were possible because the One Health space was formed by putting together a small group of participants across the agencies with mutual interest in cross-disciplinary public health work. A participant claimed that the “soft” quality of this Whole-of-Government initiative enabled compromises and agreements to be easily reached even across multiple sectors:

> It actually allows us to speak our views, maybe sometimes a bit more freely than if it’s a real, you know, formal meeting where, now whatever I say represents [my agency] and so sometimes, we are able to just sound out some of the ground issues quite openly. Yeah, I would say, my sensing is that even at the higher level, up to the Director-General level, they are obviously more formal than us when they come together for our meeting, but they are not as formal as some of the meetings that I’ve seen. So, they are reasonably formal, but they also maintain quite open conversations. They can give differing views, but they always seek a consensus in the end.

This consensus-seeking process enabled the formation of semi-informal relations that some participants referred to as a process of ‘socialisation.’ Right after an inter-agency gathering that included unresolved debates about task distribution, an attendee told MA that since every participant was busy, it was difficult to prioritise tasks and make sure there was a sense of ownership for all parties. The participant said that this sense of ownership would eventually be reached, it just required what they called “socialisation.” When MA asked a different attendee what it means to ‘socialise’ a sense of ownership, they said that it was about “slowly bringing people into consensus.” For this person, it was natural for people to express disagreements when the “buy-in” was not yet clear. The process of socialisation involved a series of ongoing discussions and related conversations that led from disagreement to a compromise and eventually also a renewed sense of ownership of the One Health space that compelled these participants to proactively engage in collaborative work.

> So when you socialise, we got to think three steps ahead… because we may not have socialized the idea initially. We went straight to step four when in their world, they are like, “What’s this? Who’s this? What am I supposed to do? How come it’s me?” Yeah, those four questions, right? We need to go four steps before to make it very friendly from their point of view to neutral, almost not uncomfortable starting point. Slowly bring them to the step four. Not jump straight to [step four].

This socialisation process and development of semi-informal relationships thus co-construct each other. Through its years of socialisation, the One Health space has moved beyond formal and professional bilateral partnerships to a uniquely warm collegiality observable in the five-agency gatherings. Before workshops and meetings officially began, there was active chatter, laughter, and discussion among participants across different agencies. Such informal discussion sometimes cut across different structural levels, with senior officials mingling with junior staff. The ensuing semi-informal relations made consensus easier to achieve. According to participants, this warmth of relations took years to develop but arose primarily due to formation of the One Health space, as one public servant described:

> In the early days, each agency will kind of deal with their own issues and if there is anything that requires any other agency’s help, it’s usually quite formal. It’s through email and then followed by the meeting. And it was very specific issues of concern. And once that is done, it’s like, you know, there’s no further engagement, unless you have further engagement required with that agency. So, when we first started […] it was more a few directors coming together to talk to each other first. Like over a cup of coffee with each agency, having that agreement to talk about whether you will be keen if you want to start off this thing.

This culture of informally catching up on potential collaborations over coffee or tea during breaks eventually became a mainstay in the One Health work culture, no longer just among directors but also at all staff levels. As an interviewee put it, this culture enabled people to “convene quickly” in case “an urgent situation arises.” A senior official shared the same sentiment:

> I think most important when something happens, you need to know who’s that person you can call up. Do you have that person on your suite dial [laughs]? On your hand phone? And do you reach out? So I think that it’s a softer side of the thing of the house. It’s not like concrete.

For all participants, the development of such “softer” relations amid otherwise formal professional relations was the value of putting together these five agencies in multi-agency gatherings:

> The big group meetings also have one benefit in that you actually learn to put the faces to all the names and it really helps that it facilitates […] rather than I just send an email cold to someone, right? And they don’t know who I am […] I can now at least say like, “Oh, we met at, you know, that One Health meeting last month and, you know, I got this issue and, you know, can we have a chat about this, you know, or something?” So, I think that that is enormously helpful.

These semi-informal relations allowed for smoother coordination when it came to health emergencies such as the Covid-19 pandemic or recent oil spill, with one participant calling the government response to the former a “smooth running of gears” due to the capacity and relationships built in the One Health space. One senior official affirmed this, saying:

> There is actually high level for support that we give each other. Me to my counterpart and vice versa. You know, the counterpart when I asked them for help, they are actually quite prepared to support as well […] So I think that’s the first point, that we do have a structure in place to facilitate this working across agencies. And then the other thing, I think, is very useful is that we do know, I mean, personally, I’ll say that I do know most of my counterparts. So not only is there official structure, there’s also the personal networks that has formed over the years because we work with each other on various things like I mentioned […]. So I actually got to know [people] better or got to know first through One Health and then after I got to know [them] better as we were on some other crisis that we faced along the way.

### Deepening connections through coordinated bilateral engagements

OHO, itself consisting of participants in the One Health space, attempts to build on and develop these semi-informal relations through increased bilateral engagement with all participants to discuss points of internal debate in more detail, providing a personal touch in the One Health space. As one participant described:

> On top of that multi-group meeting, [OHO has] been doing bilateral engagement as well and identifying our needs and see how we can put our agenda in there. So that, I think, is a very fantastic strategy. I will say it is very sincere. [They] came to us and asked us about what our needs are […]. Having a mandate, putting an office where people do it full time, I think really has enabled…this kind of connection.

Bilateral meetings facilitated efficiency and streamlined multilateral discussions by clarifying and focusing agenda items. As a senior official said:

> The bilateral meeting helped because when you go to a meeting with four [other] parties and some comments would be quite specific […If we did not have bilateral meetings with the OHO], I think we’ll probably be sucked into a little bit more minor details.

With trust built through inter-agency meetings and regular semi-informal interactions, many participants expressed how easy it had been to coordinate bilateral discussions simply and efficiently with OHO. Rather than calling them meetings, most preferred to call them “catch-ups” or “chats.” One participant explained:

> At the smaller group level, I’m quite glad that I guess we are a bit less formal and we could reach out through [virtual call] and whatever. And that also explains why we have quite a number of ad-hoc meetings sometimes. But some of the meetings were just very focused. It’s very short. We just iron out any issues. And then I will say I appreciate everyone reaching out […] to the different stakeholders and then just asking us, “What are your issues? Do you want to talk it over a 15-minute call?” And we manage to iron quite a number of issues through that. And it’s, it’s quite efficient as well.

Over time, the “friendliness factor”—as one interviewee described it—of OHO increased and it became easier to raise such feedback in other bilateral discussions. Participants appreciated these bilateral engagements. However, some also noted that while such “friendly” approaches were helpful in building connections and a sense of ownership they risked being overly “personality-driven,” as another participant put it. This participant mentioned a degree of “reliance on personal factors” in such outreach, as “some work is dependent on the ground-up efforts of vested individuals, which may not be sustainable in the longer-term or if there are changes in personnel.” This suggested that relational and formal governance mechanisms must be balanced to ensure sustainability.

## DISCUSSION

### Socialising multisectoral governance

This study found that developing multisectoral One Health governance in Singapore was predicated on socialising ‘semi-informal’ cross-institutional relationships and ownership. In this context, multisectoral One Health governance was initiated through formal structures but enacted through social networks. The social was thus central to the act of governance, which was “grounded in interaction” [26]. The structural formation of Singapore’s multisectoral One Health space thus co-constitutes the social networks crucial to national health governance.

Rotating chairmanship fostered these social networks, giving each of the five partner agencies opportunities to chair meetings and set agendas. Based on Singapore’s public service culture of rotating personnel across agencies [27, 28], this system encouraged shared responsibility, leadership experience, and adaptation to differing work cultures. While beneficial, the dispersion of responsibility in this rotating structure also raised concerns – expressed by some participants – that no single agency holds clear leadership responsibility. OHO’s creation as secretariat helps address this tension, by coordinating across rotating chairs. Centralising One Health governance under CDA – a human infectious disease-focused body - is seen as essential yet potentially limiting. Human health is widely seen as a necessary structural centre of One Health [29], yet the infectious diseases and pandemics focus raises questions about operational attention to non-communicable and non-human aspects of One Health [30]. Nonetheless, participants expressed confidence that Singapore’s One Health governance will enable consensus on priorities.

These priorities are determined through a consensus-building process participants call “socialisation,” involving multiple discussions and meetings among public servants to cultivate “semi-informal relations.” These relations maintain professional boundaries while enabling informal exchanges through texts, calls, coffee chats, and social activities. They facilitate rapid and easy knowledge transfer that has proven useful during public health emergencies requiring multisectoral coordination, affirming that social interaction is foundational to One Health governance [26]. These semi-informal relations support OHO-led bilateral engagements to resolve disagreements, follow up actions, and clarify and disseminate information across governance levels. Participants described this dependence on social relations as key to success but noted the challenge of maintaining momentum with recurring personnel rotations.

Multisectoral governance in Singapore’s One Health space is thus predicated on socialising its stakeholders. Relatedly, global literature gaps suggest the need to examine not only institutional health governance structures but also focus on individual actors influencing and driving multisectoral collaboration [31]. Building on this, our findings show that in Singapore, regular inter-agency and bilateral meetings, together with rotational chairmanship, facilitate a One Health approach that socialises public servants both with one another in formal and informal contexts and towards collective care and consensus.

### Implications

#### Formalising socialisation in health spaces

Even before CDA establishment, Singapore’s One Health approach prioritised sustainable collaboration over short-term efficiency through “semi-informal” institutionalisation of socialisation. These have proven invaluable in health crises, in which effective response depends on trust and understanding developed in “peacetime” to balance multiple stakeholder concerns.

While a multisectoral health governance model operationally based on fostering social relations between people in different agencies enhances operational effectiveness, it also raises questions about sustainability. Singapore’s rotational public service culture presents both opportunities and challenges for agenda continuity. Rotation serves as a structural safeguard against over-dependence on particular individuals and their personal relationships. However, some participants observed that the continuity of collaborative processes can be shaped by evolving priorities.

A key challenge for Singapore’s One Health space is thus formalising policies, structures, and protocols without eroding its social foundations. Bureaucratisation can routinise and dilute the ideals that make initiatives effective [32]. Yet the inter-agency commitment to embedding socialisation into formal structures reflects a determination to retain a collaborative ethos.

Based on the findings, maintaining rotational structures while institutionalising bilateral coordination and inter-agency gatherings can nurture future health leaders and sustain shared ownership. Strengthening mechanisms that encourage multisectoral socialisation may further embed intrinsic motivation for collaboration and ensure preparedness for future crises. These efforts can be codified into policies that endure beyond individual rotations.

#### Enhancing multisectoral health governance in Singapore

Singapore’s One Health approach, emphasising rotating personnel and therefore ideas across agencies facilitates recognition of trade-offs when balancing multiple interests in decision-making. The social networks and platforms described in our findings serve as conduits for frank discussions that might be more constrained in formal settings, complementing formal processes and reinforcing Singapore’s One Health governance model of consensus-building and shared ownership. While agencies maintain distinct mandates, their workstreams converge towards common goals. For instance, incorporating vector control within NEA, rather than MOH as in other countries, enables natural convergence between environmental and human disease governance. This demonstrates how institutional design complements social networks in enhancing integrated health outcomes.

Beyond One Health, other health-oriented WOG approaches indicate similar multisectoral collaboration. For example, Singapore’s anti-vaping initiative mobilises multiple bodies, including the Singapore Police Force (SPF), Central Narcotics Bureau (CNB), Health Sciences Authority (HSA), NParks, NEA, and Immigration Control Authority (ICA) [33], achieving initial successes through joint enforcement and public campaigns portraying vaping as a “toxic toy” [34]. Unlike One Health’s distributed leadership in “peacetime,” this initiative is clearly led by MOH, demonstrating how Singapore’s Whole-of-Government preparedness model is configured for crisis response.

Future research could examine how multisectoral socialisation operates in other health-oriented WOG approaches, including the roles of systematic rotation and inter-agency collaboration at a social level. Comparative analyses of such initiatives could reveal how different socialising structures enhance health outcomes. As CDA matures and new WOG efforts emerge, understanding how internal social networks interact with structural dimensions will be key to strengthening multisectoral governance for health.

### Limitations

Several limitations should be considered. Confidentiality concerns may have limited public servants’ willingness not only to participate but also to be open about their opinions, with several initially responding guardedly. Similar confidentiality concerns limited our access to One Health meetings and internal documents, narrowing observations to key gatherings that OHO and CDA considered appropriate for observation. Member checking, contextualisation through review of available documents, and interviewing a relatively large proportion of One Health participants about meetings researchers could not attend helped mitigate these limitations. Singapore’s small size and centralised governance may limit transferability and generalizability of lessons to larger or more devolved national contexts. Lastly, while not a limitation, readers should note that this study documents Singapore’s One Health structure during data collection – an initial period of OHO’s development prior to CDA’s official launch, so does not reflect any later organisational changes.

### Conclusion

Singapore’s One Health space demonstrates that effective national health governance depends on actors being well “socialised” with one another. Embedding socialisation processes within multisectoral governance is thus crucial. In Singapore, rotating chairs and personnel institutionalised social networking that supported public health and emergency response successes even before CDA’s formation. The challenge now lies in formalising these structures within a maturing One Health bureaucracy, e.g. situating the One Health approach within other WOG initiatives, without eroding its social core.

Singapore’s compact, unfederated system makes such socialisation and rotation feasible. Whether similar governance models suit larger or devolved contexts remains an open question, leaving the generalizability of the Singapore One Health model up for debate. Nonetheless, national public health agencies elsewhere can learn from Singapore by identifying and strengthening the social features of their multisectoral collaborations. Ultimately, socialisation both enables and is enabled by multisectoral governance. Understanding these reciprocal dynamics is essential to defining optimal health governance models.

## Data Availability

Due to the small sample size, redacted interview data cannot be made publicly available to ensure interviewee confidentiality.

## Acknowledgements

We would like to thank Singapore’s Communicable Diseases Agency and One Health Office for facilitating research efforts. We thank all interviewees from the five One Health partner agencies for contributing their experiences and opinions to this study. This work received technical support from the Alliance for Health Policy and Systems Research and the WHO Health Emergencies Programme.

## Author contributions

MA and NH conceptualised the study, with inputs from KL. MA collected, analysed, and interpreted data, and drafted the manuscript. PC, JC, and YN facilitated data collection and provided context. All authors contributed to interpretations and critical review. NH provided supervision and interpretation, with contributions from KL, and critical revision of the manuscript. All authors approved the version for submission.

